# Poor Accuracy of Triage Blood Pressure Measurements in a Pediatric Obesity Hypertension Clinic

**DOI:** 10.1101/2023.09.06.23295163

**Authors:** William Wen, Kevin J. Psoter, Barry S. Solomon, Elaine M. Urbina, Tammy M. Brady

**Author notes:** **Address correspondence to:** Tammy M Brady, Johns Hopkins Pediatrics, Rubenstein Child Health Building Suite 3057, 200 N Wolfe St, Baltimore, MD 21287.

## Abstract

Accurate triage blood pressure (BP) measurements are essential for clinical decision making. We investigated the agreement and diagnostic accuracy of usual triage vs. standardized BP measurements in an obesity-related hypertension clinic.

This was a retrospective study of paired triage and standardized BP measurements from patients 4-21 years old. Triage BPs are measured by a medical assistant or nurse, often by automated device. Triplicate manual BP measurements are obtained by the clinic physician. Bland-Altman analyses determined mean differences between paired triage and mean standardized BPs. GEE-based multivariable relative risk (RR) regression determined the RR of triage BP overestimation by ≥5mmHg. Overall agreement, sensitivity, specificity, positive predictive value, and negative predictive value of usual triage BP measurements identifying hypertensive BP were determined.

130 participants with 347 clinic encounters were included. Mean age was 13.3 years (standard deviation 3.94), 76% were Black race, 58% were male. Overall mean systolic and diastolic BP difference was 8.7 mmHg (95% limits on agreement: −16.66, 34.07) and 4.1 mmHg (95% limits on agreement: −18.56, 26.68), respectively. Triage systolic BP was more likely overestimated by ≥5mmHg when a large adult (RR=1.49; 95% confidence interval: 1.00, 2.21) or thigh cuff (RR=1.94; 95% confidence interval: 1.08, 3.51) was required vs. when a child/adult cuff was required. Overall agreement in identifying hypertensive BP was 57.6%. Sensitivity (52.6%), specificity (63.4%), positive predictive value (60.8%), and negative predictive value (55.3%) were low across all cuff sizes.

There was poor agreement between usual triage and standardized BP measurements, with potential for significant clinical implications.

**Clinical Trial Registration:** ReNEW Clinic Cohort Study (ReNEW), <u>NCT03816462</u>, https://clinicaltrials.gov/ct2/show/NCT03816462

## Introduction

In the past few decades, the prevalence of pediatric hypertension has risen dramatically. An estimated 1.3 million youths 12-19 years of age in the United States are currently living with hypertension, increasing their risk for left ventricular hypertrophy in childhood and cardiovascular events such as heart disease and stroke in adulthood.^1,2,3^ Hypertension in youth is often asymptomatic, and therefore diagnosis typically relies on screening. With hypertension in childhood commonly the initial sign of underlying conditions such as kidney disease, screening for hypertension is recommended at yearly preventive care visits for all children starting at 3 years of age and in infancy for at-risk children (e.g., birth <32 weeks gestation, known kidney/urologic/cardiac disease).^4,5^

In pediatric outpatient settings, blood pressure (BP) is often obtained during triage. Accurate triage measurements are essential for clinical decision making, and efficiency in taking these measurements is key for expeditious patient throughput. This can be challenging in busy settings, particularly since it can take 3.5-5 minutes to measure a single BP properly.^6^ Pediatric guidelines recommend two or more repeat measurements when the initial BP is elevated.^7^ However, this is done infrequently and clinical decisions are oftentimes based on the initial, often triage, BP measurement.^8^

In addition to repeated measurements, appropriate cuff sizing based on mid-arm circumference is required to ensure accurate readings. Significant measurement inaccuracy results when patients are mis-cuffed; discrepancies are greatest when a too-small cuff is applied for measurement [up to ∼20 mmHg systolic BP (SBP) overestimation], compared to when a too-large cuff is applied (up to ∼7 mmHg SBP underestimation), and this can result in over- or underdiagnosis of hypertension.^9^

In this study we aimed to determine the agreement and diagnostic accuracy of triage BP measurements in a vulnerable population of pediatric patients with obesity who were referred to a multidisciplinary clinic for evaluation of elevated BP. Our primary aim was to determine the agreement between usual triage and standardized BP measurements which were obtained manually according to clinical practice guidelines (CPG). We hypothesized that there would be differences between these measures and that performance would be poorer in those requiring larger BP cuff sizes. To assess the potential clinical relevance of BP measurement differences, our secondary aim was to determine the diagnostic accuracy of triage measurements in identifying hypertensive BP.

## Methods

### Study Population

The ReNEW Clinic Cohort Study (NCT03816462) is an observational cohort study designed to investigate characteristics and outcomes of pediatric patients with obesity and elevated BP. Consecutive patients with overweight/obesity and elevated BP who were referred to the ReNEW Clinic, an urban pediatric obesity-related hypertension clinic at Johns Hopkins University are invited to join the study; those who agree provide consent if ≥18 years or assent along with parent/guardian consent if <18 years. Clinic patients with elevated BPs are invited to follow up every 3-6 months while those with normal BPs are invited for follow up at 6 months. Enrollment into the ReNEW Clinic Cohort began in January 2015 and is ongoing. This study utilizes data collected between January 2015 and June 2022. The Institutional Review Board at Johns Hopkins University School of Medicine approved of this study.

### Blood Pressure Measurement

During each visit, BP is measured initially in triage predominantly with an automated device by a medical assistant or nurse, with one triage measurement recorded in the electronic medical record (EMR). After triage, patients are brought to a clinic room where BPs are measured again in triplicate via manual auscultation by the same physician (TMB) using a calibrated aneroid sphygmomanometer according to CPG.^7^ Cuff size is determined by measured mid-upper arm circumference (20-26 cm for child, 25-34 cm for adult, 32-43 cm for large adult, and 40-55 cm for thigh cuffs) and both are recorded in the EMR. Due to the limited number of participants in whom a child cuff was appropriate (n=3), child/adult cuff was combined into one category. Blood pressures were measured in the right arm (except when there was a compelling reason to use the left arm (e.g. continuous glucose monitor placement)), with the patient’s arm, back, and feet supported in a seated position. Three measurements were taken 30 seconds apart after the patient had rested for 5 minutes. The average of these triplicate measurements was used as the composite manual BP measurement for the visit.

Participants were considered to have hypertensive BP if their average manual BP was ≥ age-sex-height specific 95^th^ percentile when <13 years of age or ≥130/80 mmHg when ≥13 years of age. If the 130/80 mmHg threshold was <95^th^ percentile value for youth <13 years of age, then 130/80 mmHg was used to define a hypertensive BP.^7^

### Demographic and Clinical Characteristics

Data were abstracted from clinical forms and the EMR. Demographic data included self-reported race and biological sex. Socioeconomic status was ascertained using neighborhood-level socioeconomic indicators based on residential address. We included the most recent American Community Survey’s 5-year estimate at the block group level regarding the percent of the population ≥25 years of age with more than a high school degree; percent of the population ≥16 years of age and unemployed; percent of families with an income under the federal poverty level or receiving public assistance; and the area deprivation index.^10,11^

Anthropometric measurements were obtained by clinic staff which included height measured to the nearest 0.1 cm using a wall-mounted stadiometer and weight measured to the nearest 0.1 kg using a digital scale. Body mass index (BMI) was calculated as weight in kg/(height in m)^2^. BMI percentile and BMI z-score were calculated based on data from the Centers for Disease Control and Prevention.^12^

Presence/absence of hypertensive symptoms, medications, and cardiovascular disease risk factors were determined and collected based on review of clinic notes, and number of co-morbid cardiovascular disease risk factors was calculated by summing together instances of congenital heart disease, hypercholesterolemia, obstructive sleep apnea, and diabetes. The mean, median, and range of both total visits and follow-up visits per participant were also calculated.

### Analyses

Demographic and clinical characteristics of participants at their first visit in which triage and standardized BP measurements were recorded were summarized and compared across cuff sizes using analysis of variance (ANOVA) and Chi square or Fisher exact tests for continuous and categorical variables, respectively. Bland-Altman analyses determined the mean differences and 95% limits of agreement (LOA) between the paired triage and standardized BP measurements while accounting for the repeated measures during follow-up. Wald tests for trend using linear regression based on generalized estimating equations (GEE) were used to assess BP differences across cuff size.

The probability of having a SBP and DBP difference ≥5mmHg between concurrent triage and standardized BP at any visit was estimated using univariate and multivariable relative risk regression and fitted using a GEE-based Poisson model with robust standard errors given the non-rare (i.e. >20%) nature of the outcome.^13^ A threshold of 5 mmHg over-or-underestimation was chosen because the difference between the 90^th^ (normal) and 95^th^ percentile (hypertensive) BP across all age-sex-height strata is primarily 3-5 mmHg.^7^ Differences in risk of overestimation of SBP and DBP were compared across cuff sizes and stratified by sex using similar procedures. Multivariable models included the following *a priori* identified covariates: age, sex, BMI, height, and neighborhood poverty level (percentage of families below poverty level in each participant’s residential census tract). We repeated these analyses to determine the relative risk (RR) of underestimating BP by ≥5mmHg and of a composite outcome of either over- or underestimation by ≥5mmHg. Results of these regression models are presented as RR with corresponding 95% confidence intervals (CIs).

Next, we determined the diagnostic performance of triage measurements for identifying hypertensive BP, using the mean of the concurrent physician obtained SBP and DBP measurement at each encounter as the reference standard for hypertensive BP diagnosis. Overall agreement, sensitivity, specificity, positive predictive value (PPV) and negative predictive value (NPV), which were estimated using GEE-based logistic regression, summarized the diagnostic performance of triage measurements overall and when stratified by cuff size.^14,15^ A two-sided P value of <0.05 was considered statistically significant. Analyses were conducted using STATA Version 16.1 (StataCorp, College Station, TX) and R Statistical Software (v4.1.2; R Core Team 2021).

## Results

### Patient population

A total of 130 participants comprised the study population and contributed 347 unique clinic encounters in which paired triage and standardized BP measurements were collected. On average each participant contributed 2.7 encounters, with participants having between 1 and 13 visits within the study period. The mean age at baseline was 13.3 years (SD= 3.94) and the majority of participants were Black race (76.2%) and male (57.7%) with 42.3% having reported at least one additional cardiovascular comorbidity (Table 1). Participants’ age, BMI, and arm circumference significantly differed between cuff sizes, increasing with each incrementally larger cuff size. Participants in whom a thigh cuff was used at their initial visit had the highest prevalence of antihypertensive medication use (65%).

**Table 1:** Distribution of characteristics, by cuff size, of pediatric patients with obesity at their first clinic visit with paired triage and standardized BP measures.

| Characteristics | Overall<br>(n= 130) | Child &<br>Adult<br>(n= 50) | Large Adult<br>(n= 54) | Thigh<br>(n= 26) | P value* |
| --- | --- | --- | --- | --- | --- |
| Age (years), mean (SD) | 13.3 (3.94) | 10.7 (4.01) | 14.2 (2.82) | 16.4 (2.53) | **<0.001** |
| Sex, n (%) |  |  |  |  | 0.899 |
| Male | 75 (57.7) | 29 (58.0) | 32 (59.3) | 14 (53.9) |  |
| Female | 55 (42.3) | 21 (42.0) | 22 (40.7) | 12 (46.2) |  |
| Race, n (%) |  |  |  |  | 0.478 |
| Black | 99 (76.2) | 35 (70.0) | 42 (77.8) | 22 (84.6) |  |
| White | 20 (15.4) | 11 (22.0) | 6 (11.1) | 3 (11.5) |  |
| Other | 11 (8.5) | 4 (8.0) | 6 (11.1) | 1 (3.9) |  |
| Ethnicity, n(%) |  |  |  |  | 0.285 |
| Hispanic | 8 (6.4) | 5 (10.2) | 3 (5.9) | 0 (0) |  |
| Non-Hispanic | 118 (93.7) | 44 (89.8) | 48 (94.1) | 26 (100) |  |
| % of population with less than high school education<sup>†</sup>, mean(SD) | 15.3 (10.68) | 14.2 (10.11) | 14.5 (9.67) | 19.1 (13.05) | 0.122 |
| % of population unemployed<sup>†</sup>, mean(SD) | 11.1 (9.75) | 9.3 (9.17) | 11.6 (9.29) | 13.4 (11.38) | 0.191 |
| % of households with public assistance<sup>†</sup>, mean(SD) | 5.6 (5.90) | 5.7 (5.97) | 5.4 (5.94) | 6.0 (5.87) | 0.890 |
| % of families below poverty level<sup>†</sup>, mean(SD) | 17.0 (15.47) | 17.0 (14.46) | 16.0 (15.61) | 19.0 (17.35) | 0.719 |
| Area deprivation index<sup>†</sup>, mean(SD) | 61.3 (25.05) | 61.3 (24.04) | 61.1 (26.27) | 62.0 (25.38) | 0.988 |
| BMI (kg/m<sup>2</sup>), mean (SD) | 38.8 (9.08) | 31.8 (5.44) | 40.9 (7.50) | 47.6 (7.76) | **<0.001** |
| BMI percentile, mean (SD)<sup>‡</sup> | 99.0 (1.07) | 98.6 (1.49) | 99.2 (0.51) | 99.6 (0.50) | **0.001** |
| BMI z score, mean (SD)<sup>‡</sup> | 2.6 (0.40) | 2.5 (0.51) | 2.6 (0.27) | 2.8 (0.26) | **0.007** |
| Mid-arm circumference (cm), mean (SD) | 37.2 (6.78) | 31.6 (4.15) | 39.8 (3.77) | 46.9 (3.81) | **<0.001** |
| Standardized systolic blood pressure<sup>§</sup> (mmHg), mean (SD) | 123.3 (13.93) | 120.9 (14.04) | 123.3 (11.88) | 128.1 (16.71) | 0.095 |
| Triage systolic blood pressure (mmHg), mean (SD) | 134.3 (13.98) | 129.9 (13.52) | 135.1 (12.12) | 141.2 (15.79) | **0.003** |
| Standardized diastolic blood pressure<sup>§</sup> (mmHg), mean (SD) | 65.7 (11.28) | 65.2 (10.11) | 65.9 (11.42) | 66.5 (13.39) | 0.881 |
| Triage diastolic blood pressure (mmHg), mean (SD) | 71.2 (9.42) | 68.3 (7.93) | 71.4 (8.98) | 76.2 (10.99) | **0.002** |
| Experiencing symptoms, n(%) | 57 (43.9) | 20 (40.0) | 23 (42.6) | 14 (53.9) | 0.499 |
| Taking antihypertensive medications, n(%) | 47 (36.2) | 15 (30.0) | 15 (27.8) | 17 (65.4) | **0.003** |
| Number of additional CVD risk factors, mean (SD)<sup>#</sup> | 0.5 (0.67) | 0.4 (0.61) | 0.5 (0.72) | 0.6 (0.70) | 0.557 |
| Number of additional CVD risk factors, n(%)<sup>#</sup> |  |  |  |  | 0.268 |
| 0 | 75 (57.7) | 30 (60.0) | 32 (59.3) | 13 (50.0) |  |
| 1 | 46 (35.4) | 19 (38.0) | 17 (31.5) | 10 (38.5) |  |
| 2 | 7 (5.4) | 0 (0) | 4 (7.4) | 3 (11.5) |  |
| 3 | 2 (1.5) | 1 (2.0) | 1 (1.9) | 0 (0) |  |
| Number of visits, mean (SD) | 2.7 (2.57) | 2.6 (2.47) | 2.6 (2.50) | 2.8 (2.96) | 0.927 |
| Number of visits, median (25<sup>th</sup>, 75<sup>th</sup> percentiles) | 2 (1, 3) | 2 (1, 3) | 2 (1, 3) | 2 (1, 3) | 0.907 |
| Number of visits, median (min, max) | 2 (1, 13) | 2 (1, 12) | 2 (1, 13) | 2 (1, 12) | 0.907 |
| Number of follow-up visits, mean (SD) | 1.7 (2.57) | 1.6 (2.47) | 1.6 (2.50) | 1.8 (2.96) | 0.927 |
| Number of follow-up visits, median (25<sup>th</sup>, 75<sup>th</sup> percentiles) | 1 (0, 2) | 1 (0, 2) | 1 (0, 2) | 1 (0, 2) | 0.907 |
| Number of follow-up visits, median (min, max) | 1 (0, 12) | 1 (0, 11) | 1 (0, 12) | 1 (0, 11) | 0.907 |

\* P values based on Student tests and Chi square or Fisher exact tests for continuous and categorical variables, respectively.
† Based on the most recent American Community Survey's 5-year estimate at the block group level
‡ BMI percentiles and z-scores were calculated according to CDC data.
§ Mean of triplicate measures
|| Self-reported symptoms include headache, blurry vision, chest pain, palpitations, lightheadedness, dizziness, nausea, vomiting, and snoring.

### Differences between Triage and Standardized BP measurements

Mean triage and standardized BP was 135/71 mmHg and 126/67 mmHg, respectively. There was an overall mean SBP difference of 8.7 mmHg (95% LOA: −16.66, 34.07 mmHg) and a mean DBP difference of 4.1 mmHg (95% LOA: −18.56, 26.68 mmHg) (Figure 1). Both triage and standardized SBP and DBP measurements increased significantly across cuff sizes, and statistically significant differences between triage and standardized measurements across cuff sizes were observed for SBP, but not for DBP (Table 2). In addition, differences between triage and standardized measurements did not differ after stratifying by sex (data not shown).

**Figure 1:**
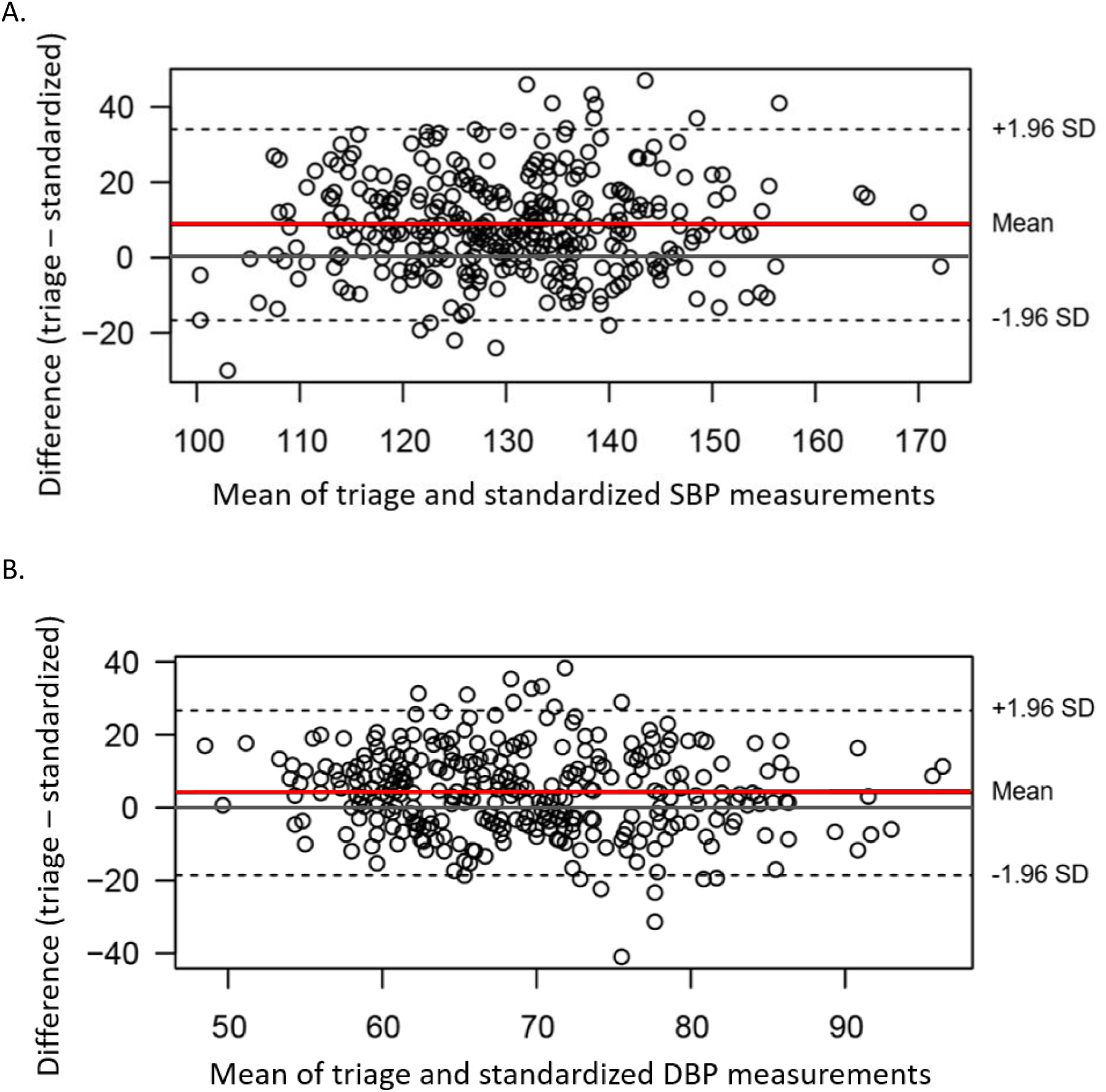
Bland-Altman plot comparing triage and standardized A) SBP and B) DBP measurements. Mean SBP difference (red line) was 8.7 mmHg and ± 1.96 SD (dashed lines) were −16.66 mmHg and 34.07 mmHg. Mean DBP difference was 4.1 mmHg and ± 1.96 SD were − 18.56 mmHg and 26.68 mmHg.

**Table 2:**
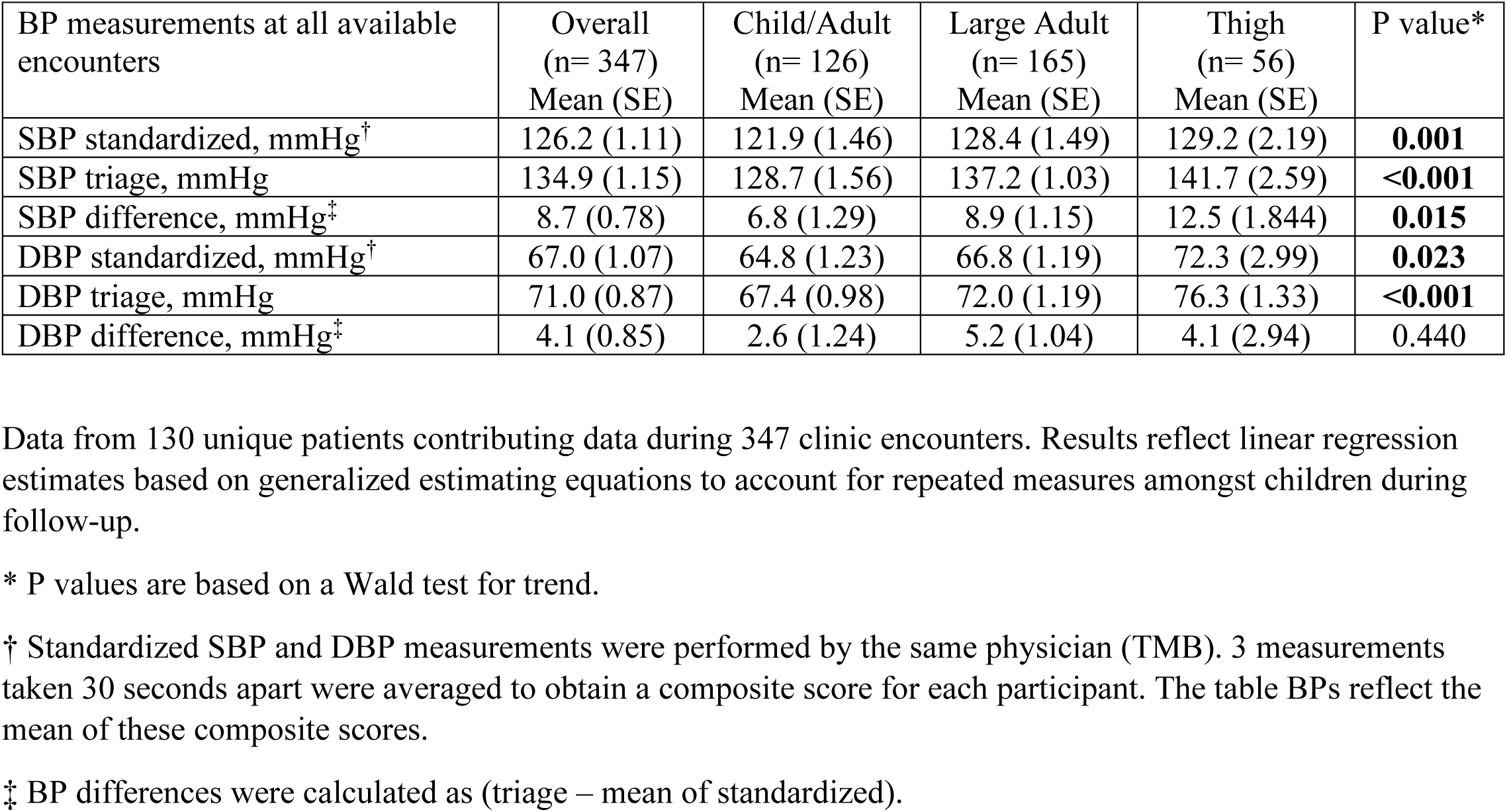
Summary of differences between all available systolic blood pressure (SBP) and diastolic blood pressure (DBP) measurements obtained in triage and by standardized physician measurement.

### Prevalence and risk of substantially different BP measurements

Of the 347 patient encounters, 257 (74.1%) had triage measurements that were different than the standardized BP by ≥5 mmHg. 211 (82%) of these encounters had a triage measurement ≥5 mmHg than the standardized measurement and 46 (18%) had triage measurements ≤5 mmHg than the standardized measurement. The estimated adjusted probability of having a substantially greater triage SBP (≥5mmHg) increased with cuff size from a low of 56.2% (95% CI: 43.2, 69.1%) for the child/adult cuff size to 65.5% (95% CI: 44.1, 86.8%) for the thigh cuff size. Similarly, the estimated adjusted probability of having a substantially greater triage DBP (≥5mmHg) increased from 44.4% (31.6, 57.2%) in the child/adult cuff size to 49.7% (26.9, 72.4%) for the thigh cuff size. Compared to SBP measurements taken with child/adult cuffs, triage BPs were significantly more likely to be overestimated for large adult (RR=1.49; 95% CI: 1.00, 2.21) and thigh cuffs (RR=1.94; 95% CI: 1.08, 3.51), while the rate of overestimation of DBP by triage measurement was similar in the large adult cuff group (RR=1.35; 95% CI: 0.86, 2.11) but higher for those requiring a thigh cuff (RR=2.01; 95% CI: 1.02, 3.97).

### Agreement and performance of triage BP for hypertension diagnosis

Overall, 52.7% of the 347 encounters had a standardized BP that was in the hypertensive range; hypertensive BP prevalence increased across cuff size (Table 3). Overall agreement of hypertensive BP classification was 57.6% and ranged from 54.7% with child/adult cuffs to 60.2% with large adult cuffs. Sensitivity (52.6%; 95% CI: 44.1, 61.2%), specificity (63.4%, 95% CI: 56.1, 70.7%), PPV (60.8%, 95% CI: 52.3, 69.2%), and NPV (55.3%, 95% CI: 47.6, 63.0%) of triage BP measurements to identify hypertensive BP were low overall and across all cuff sizes.

**Table 3:** Agreement and performance measures of recorded triage blood pressure measurements when compared to standardly obtained measurements for the identification of hypertensive blood pressure.

|  | Number of encounters<br>n (%) | Prevalence of hypertensive BP by standardized measurement* (95% CI) | Overall agreement between standardized and triage BP (95% CI) | True Positive<sup>†</sup> (# of obs) | False Positive<sup>†</sup> (# of obs) | True Negative<sup>†</sup> (# of obs) | False Negative<sup>†</sup> (# of obs) | Sensitivity of triage BP readings (95% CI) | Specificity of triage BP readings (95% CI) | PPV<sup>‡</sup> of triage BP readings (95% CI) | NPV<sup>§</sup> of triage BP readings (95% CI) |
| --- | --- | --- | --- | --- | --- | --- | --- | --- | --- | --- | --- |
| Overall | 347 (100%) | 52.7%<br>(46.4, 59.0%) | 57.6%<br>(52.5, 62.7%) | 27.7%<br>(n=96) | 17.3%<br>(n=60) | 30.0%<br>(n=104) | 25.1%<br>(n=87) | 52.6%<br>(44.1, 61.2%) | 63.4%<br>(56.1, 70.7%) | 60.8%<br>(52.3, 69.2%) | 55.3%<br>(47.6, 63.0%) |
| Cuff size |  |  |  |  |  |  |  |  |  |  |  |
| Child & Adult | 126 (36.5%) | 46.8%<br>(38.1, 55.5%) | 54.7%<br>(45.2, 64.1%) | 27.8%<br>(n=35) | 25.4%<br>(n=32) | 27.8%<br>(n=35) | 19.1%<br>(n=24) | 57.1%<br>(42.4, 71.7%) | 51.8%<br>(38.7, 64.9%) | 52.3%<br>(40.6, 64.0%) | 57.6%<br>(43.7, 71.5%) |
| Large Adult | 165 (47.6%) | 54.5%<br>(46.3, 62.8%) | 60.2%<br>(54.8, 65.5%) | 27.9%<br>(n=46) | 12.7%<br>(n=21) | 32.7%<br>(n=54) | 26.7%<br>(n=44) | 51.0%<br>(39.2, 62.8%) | 71.2%<br>(63.0, 79.4%) | 68.2%<br>(56.8, 79.7%) | 54.0%<br>(44.8, 63.1%) |
| Thigh | 56 (15.9%) | 60.7%<br>(43.7, 77.7%) | 55.3%<br>(41.3, 69.2%) | 26.8%<br>(n=15) | 12.5%<br>(n=7) | 26.8%<br>(n=15) | 33.9%<br>(n=19) | 44.0%<br>(27.0, 61.0%) | 69.1%<br>(49.5, 88.8%) | 67.7%<br>(47.0, 88.4%) | 53.1%<br>(32.4, 73.8%) |

Results reflect estimates based on generalized estimating equations to account for repeated measures amongst children during follow-up.
\* Participants were considered to have a hypertensive BP if their average triplicate physician BP measurement was ≥ age-sex-height specific 95<sup>th</sup> percentile when $<13$ years of age or $\geq 130/80$ mmHg for those 13 years of age or older. If the 130/80 mmHg threshold was lower than the 95<sup>th</sup> percentile value for youth $<13$ years of age, then 130/80 mmHg was used to define a hypertensive BP.
<sup>†</sup> Indicates how many true positives, false positives, true negatives, and false negatives would have occurred over our total 347 encounters if triage BP was solely used to identify hypertensive BP.
<sup>‡</sup> Positive predictive value
<sup>§</sup> Negative predictive value

## Discussion

In this study of pediatric patients with obesity and elevated BP referred to a multidisciplinary hypertension clinic, usual triage BP measurements had poor agreement with standardized, guideline-adherent manual BP measurements. Measurement differences were not only significant but also substantial across all cuff sizes, especially in patients requiring a thigh cuff. While sensitivity and specificity of triage measurements for identifying hypertensive BP were poor across all cuff sizes, there was a greater risk for having a triage BP measurement ≥5mmHg above the standardly obtained manual BP measurement with larger cuff sizes. These results have potentially significant implications for routine clinical care, particularly in settings where triage measurements are relied upon for clinical decision making.

Our findings are consistent with published literature. Podoll et al. found that over 74% of triage BPs obtained in a pediatric hypertension clinic at an academic medical center were higher than standardized measurements and that the majority of these triage measurements (88%) differed by ≥5mmHg.^16^ Notably, only a quarter of these participants had a BMI percentile ≥95%, whereas almost all participants in our study had BMI percentiles well above the 99^th^ percentile. They also did not report on differences by cuff size. Our study therefore fills a key knowledge gap. Further, our findings are based on updated BP percentiles and hypertension definitions and continue to demonstrate, 15 years after Podoll et al’s publication, that triage and standardized BP agreement remains low. With this discrepancy reported in hypertension specialty clinics, one could speculate that this difference may be exacerbated in general pediatric outpatient settings.

It is important to highlight that we compared a single triage BP measurement to the mean of triplicate BP measurements obtained later in the clinic session. It is well known that BP measurements decrease with repeated measurements, due to accommodation to the measurement and the regression to the mean, so it is not necessarily surprising that the standardized BPs were generally lower, and significantly so, than the single triage measurement. In fact, this phenomenon is one of the reasons the pediatric CPGs recommend repeating BPs by manual auscultation whenever the initial BP is elevated. However, due to various factors including time constraints, under-recognition of elevated readings, and lack of knowledge regarding guideline recommendations, these initial triage measurements are often not repeated and clinical decisions are made based on triage measurements. Illustrating this point, in a large, national quality improvement collaborative that included 59 pediatric practices motivated to improve the diagnosis and treatment of hypertension in a primary care setting, only 8% of patients with an elevated BP had measurements confirmed with additional BP measurements.^8^ Other studies have described similar findings: across 4 pediatric emergency departments, only 38% of patients with heightened triage BP were remeasured, and in a general pediatric outpatient clinic, only 20% with elevated readings had BP measurements repeated.^17,18^

Our results demonstrate that reliance on triage BP measurements for clinical decision making could lead to potential diagnostic and management errors. While a difference of 5 mmHg may not appear to be substantial, BP overestimation of 5 mmHg would result in 84 million adults across the globe being incorrectly classified as hypertensive.^19^ The impact on this degree of error on misclassification of hypertension in the pediatric population is still unknown, made challenging due to the reliance on percentile-based definitions for those under 13 years of age. However, the difference between the 90^th^ and 95^th^ percentile BP across all age-sex-height strata is primarily 3-5 mmHg, suggesting that even a difference of 5 mmHg can lead to drastically different diagnoses in children.^7^ If triage BP measurements were solely used to inform clinical decisions in the ReNEW clinic, 17% (60/347) of the clinical encounters would have ended with unnecessary medication initiation or escalation, labs, or imaging due to a hypertension misdiagnoses, and 25% (87/347) of encounters would have falsely reassured patients and families, resulting in a missed opportunity for early evaluation and intervention.

While not rigorously evaluated in this study, it is possible that some of the BP differences found between the triage and standardized BPs were due to measurement technique. Many studies have identified technique errors as a major cause of BP inaccuracies. In a multisite cohort study surveying for adherence to the 2017 American Academy of Pediatrics’ CPG, Rea et al. found that only 2% of patients had all BP measurement steps completed correctly.^8^ A study of 159 medical students found that on average only 4 of the 11 essential steps were completed, with adherence to rest, feet support, quiet environment without talking or cell phone use most infrequently completed.^20^ Ray et al. have reported an average of 4 errors in BP measurement technique per patient encounter, with the most common errors including lack of adequate rest (93%), poor feet positioning (48%), incorrect cuff sizing (40%), and measurement of BP over clothing (93%).^21^ We did not directly observe triage BP measurements for all 347 encounters in this study and documentation often did not include method of measurement (manual auscultation or automated). However, triage measurements by several clinic staff observed in July 2022 revealed poor adherence to the following: arm, back, and feet support; mid-arm circumference measurement to inform cuff size selection; and 3-5 minutes antecedent rest prior to measurement. This contrasts with the standardly measured BPs: patients void prior to the start of the clinic visit, three manual BP measurements are obtained after 5 minutes of rest, with proper positioning, individualized cuff size selection based on measured mid-arm circumference, and in silence with cell phones out of reach.

There are several limitations to our study. We report results from a single multidisciplinary clinic at an urban academic medical center which may not be generalizable to other settings where children and adolescents receive medical care. In addition, ABPMs (ambulatory blood pressure monitors) were not available to confirm physician obtained BP elevations. While initially incorporated systematically in the clinic evaluation for all patients, due to low rates of ABPM device return, 24-hour ABPM is now employed in the ReNEW clinic on a case-by-case basis. It is possible that some of the BP elevations noted during standardized measurement were due to the white coat effect, something we could not assess without ABPM. Although it is likely that the large discrepancy between triage and standardized BP measurements is related in part to measurement techniques, we do not have data detailing how the triage measurements were performed for the 347 encounters.

Our study also has many strengths. We present results from a robust cohort of 130 participants with 347 encounters in which triage and standardized BP measurements were obtained over the course of 7 years. The participants have a wide range of age, sex, socioeconomic status, BMI, and comorbidities. By exploring the accuracy of triage measurements in a pediatric population that is particularly vulnerable to the repercussions of inaccurate BP measurements and management, we highlight the clinical implications in a highly relevant patient population.

### Perspectives

In summary, BP screening in children is a recommended practice for early diagnosis and treatment of hypertension in childhood. In a multidisciplinary pediatric obesity-related hypertension clinic, we found that usual triage measurements differed significantly and substantially from standardized manual BP measurements, especially for patients needing larger cuff sizes. Low diagnostic accuracy in identifying hypertensive BP suggests that there may be considerable under and over-diagnoses of hypertension if triage BP are used to inform clinical decisions. Our results thus point towards the importance of adhering to proper BP measurement guidelines and the need to confirm elevated triage BP with repeated measurements done via manual auscultation according to CPGs. They also suggest that efforts to standardize triage measurements hold promise for improving patient care as well as clinic efficiency.

## Data Availability

The data that support the findings of this study are not publicly available but are available from the corresponding author upon reasonable request.

## Acknowledgements

None

## Nonstandard Abbreviations

ABPM: ambulatory blood pressure monitoring
ANOVA: analysis of variance
BMI: body mass index
BP: blood pressure
CI: confidence interval
CPG: clinical practice guidelines
DBP: diastolic blood pressure
EMR: electronic medical record
GEE: generalized estimating equations
LOA: limits of agreement
NPV: negative predictive value
PPV: positive predictive value
RR: relative risk
SBP: systolic blood pressure
SD: standard deviation

## Novelty and Relevance

### What is Relevant?

Accurate triage BP measurement is essential for clinical decision making, but many important BP measurement steps such as repeating elevated measurements and individualizing cuff size selection are omitted in busy settings, threatening the accuracy of measurements.

### What is New?

This study investigates the agreement and diagnostic accuracy of usual triage with standardized blood pressure measurements in children referred to a subspecialty obesity-related hypertension clinic. We found low agreement and poor diagnostic accuracy of triage BP measurements across all cuff sizes, but especially for those needing larger cuffs.

## Clinical Implications

Renewed emphasis on measurement technique in triage could improve patient care and throughput.

## References

1. Jackson SL, Zhang Z, Wiltz JL, et al. Hypertension Among Youths - United States, 2001-2016. MMWR Morb Mortal Wkly Rep. Jul 13 2018;67(27):758–762. doi:10.15585/mmwr.mm6727a2

2. Price JJ, Urbina EM, Carlin K, et al. Cardiovascular Risk Factors and Target Organ Damage in Adolescents: The SHIP AHOY Study. Pediatrics. Jun 1 2022;149(6)doi:10.1542/peds.2021-054201

3. Jacobs DR, Jr., Sinaiko AR, Woo JG. Childhood Risk Factors and Adult Cardiovascular Events. Reply. N Engl J Med. Aug 4 2022;387(5):473–474. doi:10.1056/NEJMc2208135

4. Hadtstein C, Schaefer F. Hypertension in children with chronic kidney disease: pathophysiology and management. Pediatr Nephrol. Mar 2008;23(3):363–71. doi:10.1007/s00467-007-0643-7

5. Gartlehner G, Vander Schaaf EB, Orr C, Kennedy SM, Clark R, Viswanathan M. Screening for Hypertension in Children and Adolescents: Updated Evidence Report and Systematic Review for the US Preventive Services Task Force. JAMA. Nov 10 2020;324(18):1884–1895. doi:10.1001/jama.2020.11119

6. Brady TM, Charleston J, Ishigami J, Miller ER, 3rd, Matsushita K, Appel LJ. Effects of Different Rest Period Durations Prior to Blood Pressure Measurement: The Best Rest Trial. Hypertension. Nov 2021;78(5):1511–1519. doi:10.1161/HYPERTENSIONAHA.121.17496

7. Flynn JT, Kaelber DC, Baker-Smith CM, et al. Clinical Practice Guideline for Screening and Management of High Blood Pressure in Children and Adolescents. Pediatrics. Sep 2017;140(3)doi:10.1542/peds.2017-1904

8. Rea CJ, Brady TM, Bundy DG, et al. Pediatrician Adherence to Guidelines for Diagnosis and Management of High Blood Pressure. J Pediatr. Mar 2022;242:12–17 e1. doi:10.1016/j.jpeds.2021.11.008

9. Ishigami J, Charleston J, Miller ER 3rd, Matsushita K, Appel LJ, Brady TM. Effects of Cuff Size on the Accuracy of Blood Pressure Readings: The Cuff(SZ) Randomized Crossover Trial. JAMA Intern Med. 2023 Aug 7:e233264. doi: 10.1001/jamainternmed.2023.3264. Epub ahead of print. PMID: 37548984; PMCID: PMC10407761.

10. Singh GK. Area deprivation and widening inequalities in US mortality, 1969-1998. Am J Public Health. Jul 2003;93(7):1137–43. doi:10.2105/ajph.93.7.1137

11. Kind AJ, Jencks S, Brock J, et al. Neighborhood socioeconomic disadvantage and 30-day rehospitalization: a retrospective cohort study. Ann Intern Med. 2014;161(11):765–774. doi:10.7326/M13-2946

12. Growth charts - clinical growth charts. Centers for Disease Control and Prevention. http://www.cdc.gov/growthcharts/clinical_charts.htm. Published June 16, 2017. Accessed January 30, 2023.

13. Zou GY, Donner A. Extension of the modified Poisson regression model to prospective studies with correlated binary data. Stat Methods Med Res. Dec 2013;22(6):661–70. doi:10.1177/0962280211427759

14. Genders TS, Spronk S, Stijnen T, Steyerberg EW, Lesaffre E, Hunink MG. Methods for calculating sensitivity and specificity of clustered data: a tutorial. Radiology. Dec 2012;265(3):910–6. doi:10.1148/radiol.12120509

15. Lim Y. A GEE approach to estimating accuracy and its confidence intervals for correlated data. Pharm Stat. Jan 2020;19(1):59–70. doi:10.1002/pst.1970

16. Podoll A, Grenier M, Croix B, Feig DI. Inaccuracy in pediatric outpatient blood pressure measurement. Pediatrics. Mar 2007;119(3):e538–43. doi:10.1542/peds.2006-1686

17. Silverman MA, Walker AR, Nicolaou DD, Bono MJ. The frequency of blood pressure measurements in children in four EDs. Am J Emerg Med. Nov 2000;18(7):784–8. doi:10.1053/ajem.2000.16311

18. Koebnick C, Mohan Y, Li X, et al. Failure to confirm high blood pressures in pediatric care-quantifying the risks of misclassification. J Clin Hypertens (Greenwich). Jan 2018;20(1):174–182. doi:10.1111/jch.13159

19. Padwal R, Campbell NRC, Weber MA, et al. The Accuracy in Measurement of Blood Pressure (AIM-BP) collaborative: Background and rationale. J Clin Hypertens (Greenwich). Dec 2019;21(12):1780–1783. doi:10.1111/jch.13735

20. Rakotz MK, Townsend RR, Yang J, et al. Medical students and measuring blood pressure: Results from the American Medical Association Blood Pressure Check Challenge. J Clin Hypertens (Greenwich). Jun 2017;19(6):614–619. doi:10.1111/jch.13018

21. Ray GM, Nawarskas JJ, Anderson JR. Blood pressure monitoring technique impacts hypertension treatment. J Gen Intern Med. Jun 2012;27(6):623–9. doi:10.1007/s11606-011-1937-9

